# Mitochondria-mediated Maternal-fetal Interactions and Consequences of Mitochondrial Dysregulation Indicate New Roles for Mitochondria in Hypertensive Pregnancies

**DOI:** 10.1101/2021.12.18.21268029

**Authors:** Contessa A. Ricci, Danielle M. Reid, Jie Sun, Donna A. Santillan, Mark K. Santillan, Styliani Goulopoulou, Nicole R. Phillips

**Affiliations:** Department of Physiology and Anatomy, University of North Texas Health Science Center, Fort Worth, Texas 76107, USA; Department of Microbiology, Immunology and Genetics, University of North Texas Health Science Center, Fort Worth, Texas 76107, USA; Department of Obstetrics and Gynecology, University of Iowa Carver College of Medicine, Iowa City, Iowa 52242, USA

## Abstract

Oxidative stress, placental mitochondrial morphological alterations, and impaired bioenergetics are associated with hypertensive disorders of pregnancy. Here we examined mitochondrial DNA mutational load in pregnant women with pregnancy-induced hypertension and reanalyzed publicly available high-throughput transcriptomic datasets from maternal and fetal tissues from normotensive and hypertensive pregnancies. Mitochondrial dysregulation was indicated by aberrant mitochondrial gene expression, and putative consequences were examined. Women with hypertensive pregnancy had elevated mitochondrial DNA mutational load. Maternal mitochondrial dysregulation in hypertensive pregnancies was associated with pathways involved in inflammation, cell death/survival, and placental development. In fetal tissues from hypertensive pregnancies, mitochondrial dysregulation was associated with increased extracellular vesicle production. Our study demonstrates mitochondria-mediated maternal-fetal interactions during healthy pregnancy and maternal mitochondrial dysregulation in hypertensive pregnancy development.

## Introduction

Impairments in mitochondria fusion/fission dynamics and oxidative phosphorylation[2, 3] are implicated in new-onset hypertensive disorders of pregnancy like gestational hypertension and preeclampsia. However, consequences of mitochondrial dysfunction in hypertensive pregnancies are not fully described, nor is the role of maternal versus fetal mitochondrial dysfunction clearly delineated. In support of the hypothesis that mitochondrial dysfunction underlies pregnancy-induced hypertension, we recently demonstrated impaired circulating cell-free mitochondrial DNA (ccf-mtDNA) dynamics in pregnant mothers with preeclampsia[4]. This finding is highly relevant to preeclampsia pathophysiology, as ccf-mtDNA is often used as a non-invasive proxy for cellular stress and systemic inflammation due to its ability to activate Toll-like receptor 9 and other DNA-sensing receptors[5, 6]. Here we examined ccf-mtDNA integrity in maternal samples from normotensive and new-onset hypertensive pregnancies. We additionally analyzed publicly available maternal and fetal high-throughput transcriptomic datasets from normotensive and new-onset hypertensive pregnancies. We hypothesized that 1) maternal ccf-mtDNA from pregnancies with hypertension would have higher mutational loads compared to normotensive pregnancies; 2) aberrant expression of mitochondrial genes (i.e., mitochondrial dysregulation) would be present in maternal and fetal tissues from pregnancies with hypertension; and 3) putative consequences of mitochondrial dysregulation in pregnancies with hypertension would mirror processes known to have roles in hypertensive pregnancy pathology.

## Results and discussion

We first quantified ccf-mtDNA mutational load in maternal buffy coat collected in the 3^rd^ trimester from 13 normotensive pregnancies and 10 pregnancies with preeclampsia. Because mitochondria are maternally inherited, maternal and fetal ccf-mtDNA contributions could not be known. Subsequently, we analyzed publicly available maternal and fetal high-throughput transcriptomic datasets. Data were accessed from a) maternal blood plasma of 9 normotensive pregnancies and 8 pregnancies with preeclampsia or gestational hypertension[7]; and b) fetal placenta of 21 normotensive pregnancies and 20 pregnancies with preeclampsia[1]. Maternal samples were collected during 1^st^, 2^nd^, and 3^rd^ trimesters, and at delivery. Fetal samples were collected at delivery. For maternal data, the original publication did not distinguish between mothers experiencing preeclampsia or gestational hypertension. Although gestational hypertension may be considered sub-clinical preeclampsia[8], we acknowledge the pathologies included here are not restricted to a specific hypertensive subtype. We collectively refer to pregnancies with preeclampsia or gestational hypertension as hypertensive pregnancies hereon.

Elevated ccf-mtDNA mutational loads were observed in mothers with hypertensive pregnancies (Figure 1A), but all detected differentially expressed mitochondrial genes (mtDEGs) between normotensive and hypertensive pregnancies (maternal and fetal) were nuclear-encoded (Supplementary Table S2). The absence of mitochondria-encoded mtDEGs despite increased ccf-mtDNA mutations may reflect constraints on mitochondrial function as an independent organelle. Mitochondria-encoded genes are limited (Complex I/III/IV/V subunits and rRNAs/tRNAs for mitochondrial translation[9-11]) and are less regulated than nuclear-encoded mitochondrial genes[9, 12, 13]. Additionally, mitochondrial fidelity is largely driven by nuclear-encoded genes (e.g., master regulators of mitochondrial biogenesis[10] and dynamics[10, 11]). Therefore, compensation may easily occur for ccf-mtDNA mutations. Mitochondrial dysregulation was present in hypertensive maternal and fetal tissues, where 30 mtDEGs were detected in maternal blood (Figure 1B), and 9 were detected in fetal placenta (Figure 1C). Only 2 mtDEGs were shared between maternal gestational ages: *MRPL38* (1^st^ trimester and at delivery) and *BCL2L1* (3^rd^ trimester and at delivery). Paired with increased ccf-mtDNA mutational load, this finding suggests generalized mitochondrial dysregulation in hypertensive mothers (versus a specific point of breakdown) and may not be specific to pregnancy. Though fetal gene expression was not tracked through gestation, it is worth noting that no mtDEGs were shared between maternal and fetal datasets. However, because datasets originated from separate studies and different tissues, further implication should be observed with caution.

**Figure 1.**
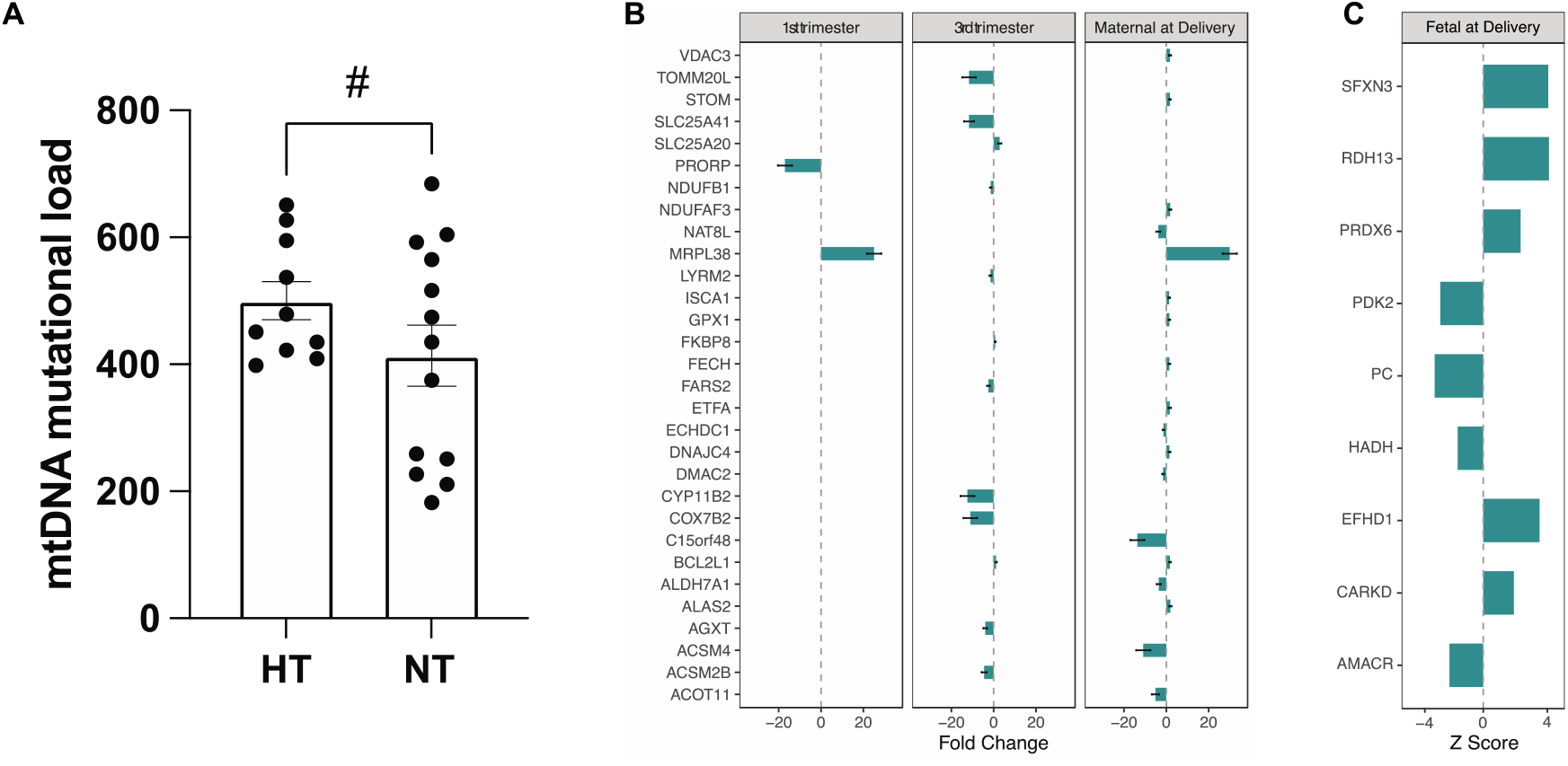
Maternal mitochondrial DNA mutational load and mtDEGs in maternal and fetal tissues. A) maternal ccf-mtDNA mutational load; “HT”: hypertensive pregnancies (n = 10); “NT”: normotensive pregnancies (n = 13); “#”: trending on significance (upaired t-test, P-value = 0.08); medium effect size observed (Cohen’s *d* = 0.60), indicating biological significance. B) maternal mtDEGs in blood plasma throughout gestation and at delivery (no mtDEGs were detected in 2^nd^ trimester). C) fetal mtDEGs in placenta at delivery; P-adjusted values of significance and expression directions provided in the original publication were converted to z-scores; error bars not present because standard error was not reported in original publication[1]

Of the 14 maternal mtDEGs specific to gestation, 5 displayed changes in the expression of genes within their interaction networks (hereon, mitochondrial interaction genes) that were differentially affected by gestational age and condition (Figure 2A; Supplementary Table S3). Of these, all but 1 (*SLC25A41* interaction genes) displayed decreased expression through gestation in hypertensive mothers. All 5 significant gene sets were primarily enriched for mitochondrial and metabolic homeostatic processes (Gene Ontology enrichment[14]; Supplementary File 3).

**Figure 2.**
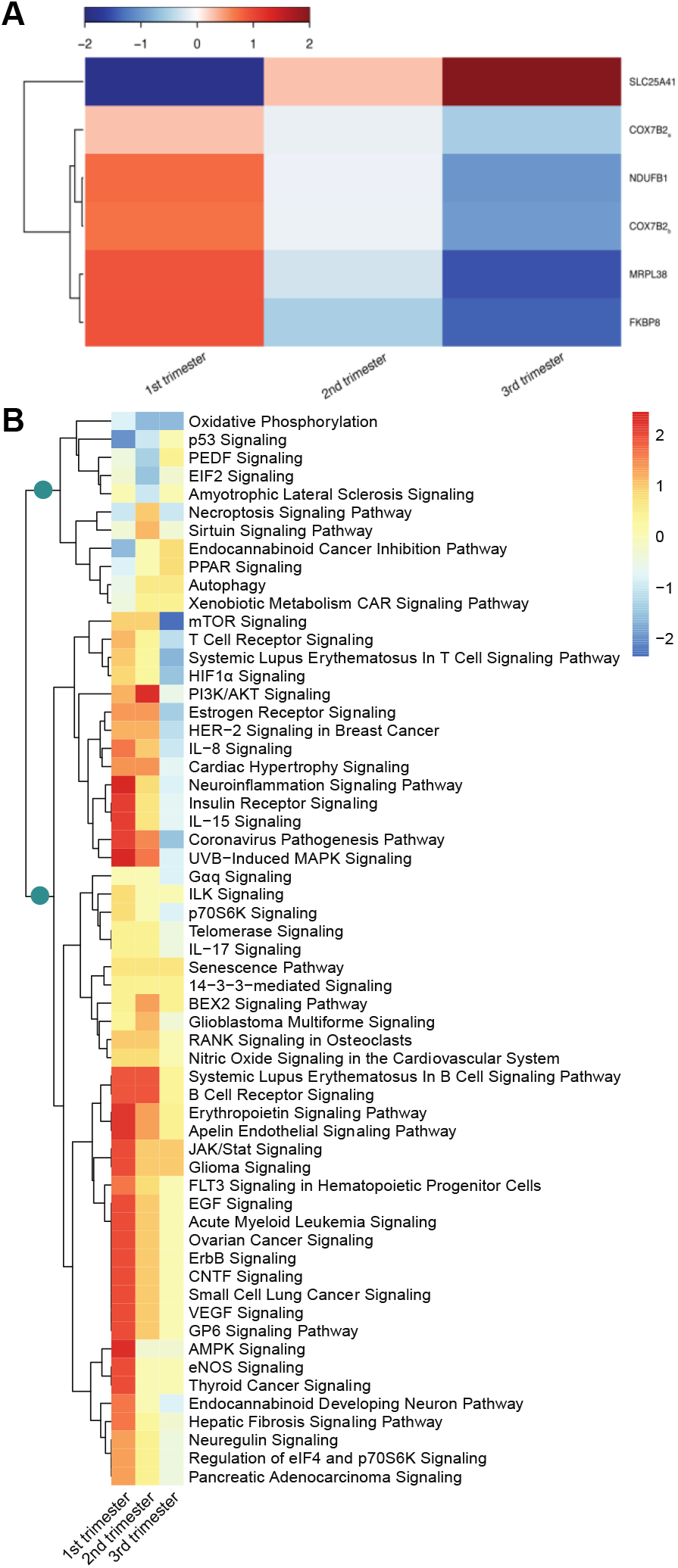
Maternal mitochondrial interaction gene expression through gestation. A) mitochondrial interaction gene sets significantly affected by time and gestational age; subscript for *COX7B2* indicate 2 separate but significant gene expression trends were detected. B) hierarchical clustering of differentially regulated pathways in *MRPL38* and *FKBP8* interaction genes; teal circles represent groupings of cell death/survival pathways and immune system pathways.

*MRPL38* and *FKBP8* interaction genes were additionally enriched for immune system and cell death/survival pathways (Figure 2B). Pathway analysis[15] (Ingenuity Pathway Analysis, QIAGEN Inc., https://www.qiagenbioinformatics.com/products/ingenuity-pathway-analysis) of *FKBP8* and *MRPL38* interaction genes in hypertensive mothers showed hierarchical clustering into 2 main branches: a smaller group overrepresented by cell death/survival pathways (approximately 45% of 11 total pathways; Supplementary Table S4) displaying downregulation in early pregnancy that transitioned to upregulation in late pregnancy; and a larger group overrepresented by immune system pathways (inflammation in particular; approximately 35% of 48 total pathways; Supplementary Table S4) displaying upregulation in early pregnancy that was either attenuated or transitioned to downregulation in late pregnancy. This opposing pattern may be a mitigation response to overactivation of inflammatory processes. Increased ccf-mtDNA mutations may also contribute here, as mitochondrial DNA **A** harboring oxidative lesions is directly linked to mutagenesis/mutational load[16-18] and is more immunogenic[18-21]. Because damage may occur very early on, or mothers may enter pregnancy with unknown underlying pathologies, these results highlight the need for early detection of risk for hypertensive pregnancy and organ damage to manage and/or improve maternal and fetal outcomes. Ccf-mtDNA may be fruitful, as ccf-mtDNA mutational load earlier in pregnancy may be more pronounced than in the 3^rd^ trimester.

Vascular endothelial growth factor (VEGF) and peroxisome proliferator-activated receptor (PPAR) signaling, key molecules implicated in preeclampsia, were present in *FKBP8* and *MRPL38* interaction gene pathways. PPARs (PPARγ in particular) are associated with placental development and differentiation[22]. Here, early hypertensive pregnancy had downregulated PPAR signaling. Because the PPARγ protein is expected to be consistently lower in serum from mothers with preeclampsia[22, 23], the observed transition to upregulated PPAR signaling in late hypertensive pregnancy may reflect pathway crosstalk or compensatory processes that ensure offspring survival.

VEGF is involved in placental vasculogenesis[24]. It is implicated in preeclampsia via effects on maternal endothelial function when VEGF signaling is aberrantly antagonized by the soluble form of placental fms-like tyrosine kinase 1 (sFLT1) [25, 26]. Importantly, though *FLT1* is not a mitochondrial interaction gene, it was upregulated at delivery in the hypertensive fetal placentas examined here[1]. Because sFLT1 is only expected to be upregulated in late pregnancy in preeclampsia (and not different during early pregnancy)[27], antagonism by circulating sFLT1 likely only contributes to the attenuated maternal VEGF signaling late in pregnancy observed here. The upregulated VEGF signaling in early hypertensive pregnancy thus supports the angiogenic imbalance hypothesis[28], where dysregulated (versus impaired) angiogenesis plays a major pathogenic role in pregnancy-induced hypertension. Although we did not identify differential expression of *VEGF* in maternal plasma, VEGF upregulation has been observed in preeclamptic decidua[25], and VEGF overexpression in pregnant mouse endometrium produces preeclampsia-like signs[25].

At delivery, 18 maternal mtDEGs were detected but only 37 mitochondrial interaction genes were simultaneously differentially expressed (Supplementary File 5). Likewise, the 9 mtDEGs detected in fetal placentas were associated with only 13 differentially expressed mitochondrial interaction genes (Supplementary File 5). Maternal mitochondrial interaction genes were enriched (Gene Ontology enrichment[14]) for approximately 9 main biological processes in hypertensive mothers (Figure 3A; Supplementary File 5) but lacked a unifying theme. Fetal mitochondrial interaction genes in hypertensive pregnancies were only enriched for metabolic biological processes (Figure 3A; Supplementary File 5). These data suggest mitochondria-mediated biological processes at delivery are not involved in hypertension, or that mitochondria-mediated hypertensive processes are alleviated. However, fetal mitochondrial interaction genes were enriched for 21 cellular components that were almost exclusively indicative of cellular secretion and extracellular vesicle (EV) release (Figure 3B; Supplementary File 5). Conversely, cellular components differentially enriched among mitochondrial interaction genes in hypertensive mothers also lacked a unifying theme and shared only a single term (vesicles) with fetal enrichments (Figure 3B; Supplementary File 5). Though the lack of shared enrichments between maternal and fetal tissues is likely an artifact of sample tissue origin, it may also reflect the fundamentally different biological priorities at delivery between mothers and neonates.

**Figure 3.**
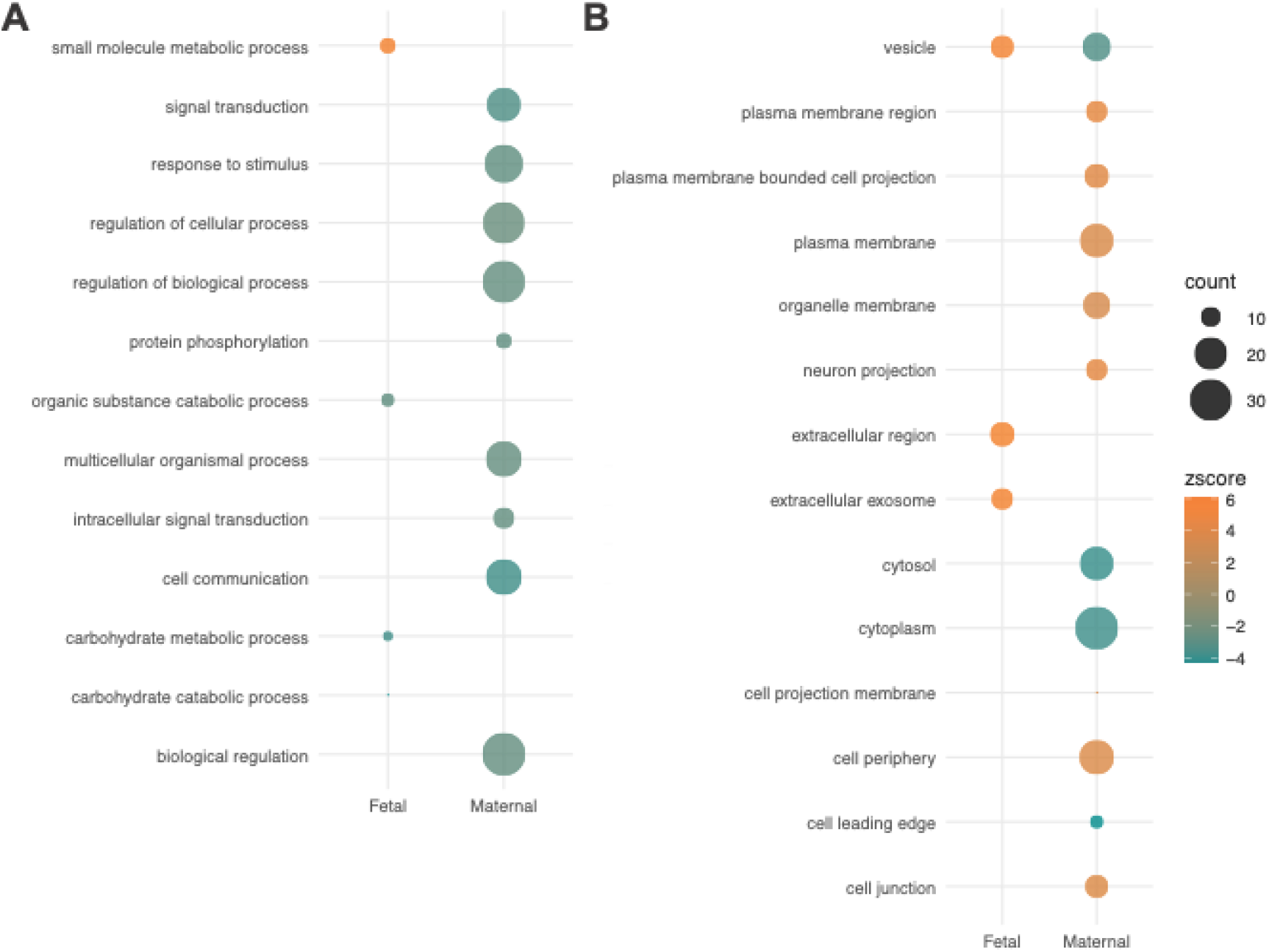
Expression patterns and functional enrichment analysis of maternal and fetal mitochondrial interaction genesat delivery. A) biological process enrichments for hypertensive maternal and fetal tissues at delivery. B) cellular component enrichments for hypertensive maternal and fetal tissues at delivery. Terms condensed for plotting purposes if genes comprising the term shared ≥ 99% overlap. “Count” represents number of genes comprising term.

The prominent signal for EV production in hypertensive fetal placentas is notable. As placenta-derived EVs are more abundant in pregnancies with preeclampsia[29], our findings suggest a direct consequence for fetal mitochondrial dysregulation in cell-cell communication, and possible delivery of placental factors that can affect maternal physiology. Further, our work has shown ccf-mtDNA primarily exist in a membrane-bound state in both normotensive mothers and mothers with preeclampsia[4], and others have shown the presence of sFLT1 in placenta-derived EVs[30, 31]. If this pattern is present throughout pregnancy, it may implicate fetal inheritance of mitochondrial dysregulation as an important contributor to pregnancy-induced hypertension, particularly as it pertains to placental function and communication between mother and fetus.

## Conclusions and perspectives

We provide evidence that mitochondrial dysregulation contributes to pregnancy-induced hypertension. We demonstrate increased ccf-mtDNA mutational load associated with hypertensive pregnancy, aberrant expression of mitochondrial genes, and aberrant expression of mitochondrial interaction genes, through pregnancy and at delivery. We connect pregnancy-specific mitochondrial dysregulation with established preeclampsia-associated processes (VEGF and PPAR signaling) and inflammation. We also show fetal placental mitochondrial dysregulation at delivery is associated with increased EV production. Though this may be specific to delivery, we hypothesize that increased placental EV production observed in other studies during preeclamptic pregnancy may also be mitochondria-mediated. Following our findings, we propose a two-pronged theoretical model (Figure 4) where maternal mitochondrial dysregulation promotes the uncontrolled, over-inflammatory state in pregnancy-induced hypertension through angiogenic imbalance and ccf-mtDNA-mediated immunogenicity. Simultaneously, inherited mitochondrial dysregulation leads to increased EV release during placental and fetal development, which then modulates systemic maternal physiology via introduction of pro-inflammatory, placenta-derived factors into maternal circulation.

**Figure 4.**
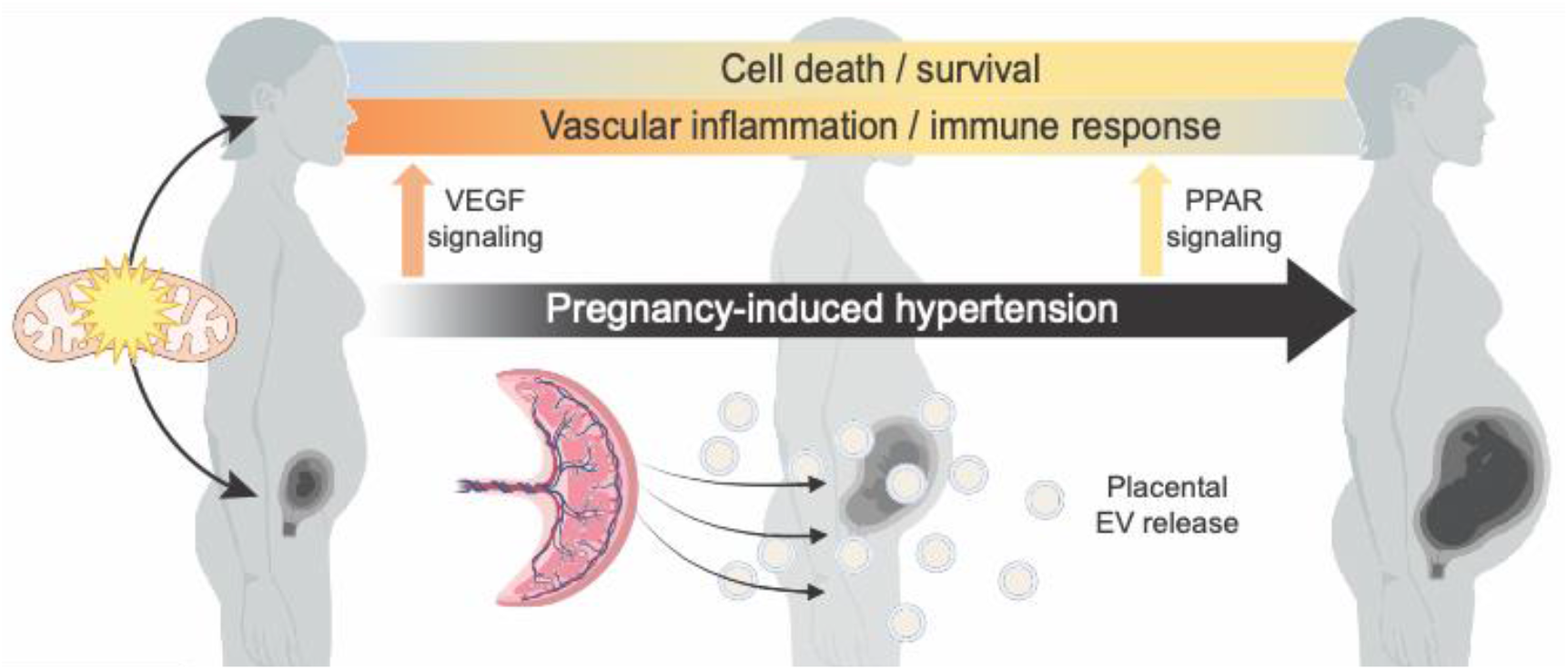
Theoretical model depicting two-pronged effects of mitochondrial dysregulation in pregnancy-induced hypertension development. In mothers with pregnancy-induced hypertension, mitochondrial dysregulation leads to angiogenic imbalance and differential regulation of cell death/survival pathways and inflammatory/immune response pathways, in addition to dysregulation of VEGF (increased in early gestation) and PPAR (increased in late gestation) signaling. In the developing fetus, inheritance of mitochondrial dysregulation affects maternal-fetal communication through increased extracellular vesicle (EV) release from the placenta.

## Data Availability

All data in the present study are publicly available via:
NCBI Gene Expression Omnibus:
GEO accession GSE154377
GSE accession GSE114691
GitHub:
https://github.com/contessaricci/mito_dysfunction_PIH

## Acknowledgements

Computational and Analytics resources were provided by the North Texas Scientific Computing, a division under the of office of the Chief Information Officer for UNT and UNT System. This research was supported in part by the National Institutes of Health (R01HL0146562; R01HD089940), National Institutes On Minority Health And Health Disparities of the National Institutes of Health (U54MD006882), American Heart Association (18SCG34350001, 15SFRN 23730000, 15 SFRN 23480000), the University of Iowa Carver College of Medicine, and the National Center for Advancing Translational Sciences of the National Institutes of Health (UL1TR002537, UL1TR002537-S1). The content is solely the responsibility of the authors and does not necessarily represent the official views of funders.

## Materials and methods

### Mitochondrial DNA mutational load

Mutational load analysis was carried out using whole mitochondrial genome amplification followed by deep Illumina-based next generation sequencing. Subject characteristics and sample collection are detailed in [4]. Sample collection occurred during the 3^rd^ trimester (28 – 41 weeks of gestation). Whole mitochondrial DNA was amplified via REPLI-g Human Mitochondrial DNA kit (Qiagen, Venlo, Netherlands) following the manufacturer protocol. The method of amplification uses phi29 polymerase-based rolling circle and multiple displacement amplification. This method was selected to effectively enrich for the mitochondrial genome as opposed to nuclear DNA, as a means for sufficient coverage for subsequent whole genome sequencing targeting mtDNA. The sequencing library for each sample was prepared via the Nextera XT DNA Library Preparation kit (Illumina, San Diego, CA) following the manufacturer protocol for multiplexing. The sequencing library was sequenced on the NextSeq 550 Sequencer (Illumina) platform. Raw sequenced reads were aligned to the human mitochondrial reference genome hg38 via BWA-MEM (v0.7.17) using the default parameter for mapping to produce corresponding SAM files[32]. The generated SAM files were processed with SAMtools (v.1.9) to produce BAM files that were sorted, indexed, and statistically assessed by coordinate[33]. Reads within the generated BAM files were assigned to a single new read-group through Picard via the AddOrReplaceReadGroups tool (http://broadinstitute.github.io/picard). The resultant single read-group BAM files were further processed to remove duplicate reads using the GATK4 Spark application of the Picard tool MarkDuplicates[34]. SAMtools (v.1.9) was used to index the reads followed by somatic variant calling via GATK4 Mutect2 utilizing the mitochondrial mode and excluding read orientation base qualities below 30[34, 35]. Variants confirmed in both read directions were tallied. The data were tested and confirmed for normality (Supplementary Figure 1), and an unpaired t-test (one-sided) was used to test the hypothesis that mothers with PE have elevated mtDNA mutational load. T-test and graphs were generated using Prism v.9.1.1. Effect size was determined by Cohen’s *d* estimation[36-38] using the “cohen.d” function in EffSize[39] package available for R, where small effect sizes are a Cohen’s *d* of approximately 0.2, medium effect sizes are a Cohen’s *d* of approximately 0.5, and large-effect sizes are a Cohen’s *d* of approximately 0.8[38, 39].

### Transcriptomics data description

Two high-throughput sequencing transcriptomic datasets were obtained through NCBI’s Gene Expression Omnibus[40]. All code and working files are publicly accessible via Github (https://github.com/contessaricci/mito_dysfunction_PIH). Longitudinal data from peripheral blood plasma of 9 mothers with normotensive pregnancies and 8 mothers with pregnancies complicated with preeclampsia and/or gestational hypertension (GEO accession GSE154377; [7]) were used to investigate maternal dynamics of aberrant mitochondrial gene expression. Patient-specific data can be found in original publication[7]. These data were sequenced on the Illumina HiSeq 4000 platform. Data were collected at four time points throughout gestation: at the end of the first trimester (referred to as 1^st^ trimester, collected between 12–17 weeks of gestation), at the end of the 2^nd^ trimester (referred to as 2^nd^ trimester, collected between 18–22 weeks of gestation), at the end of the 3^rd^ trimester (referred to as 3^rd^ trimester, collected between 35–37 weeks of gestation), and at delivery (time of collection unspecified). Data from placental samples collected within 30 minutes of delivery were used from 21 normotensive pregnancies and 20 pregnancies complicated by early-onset preeclampsia (GSE accession GSE114691; [1]). Placental samples were collected from two central and two peripheral locations, pooled, and sequenced on the Illumina HiSeq 2000 platform. Patient-specific data can be found in original publication[1]. Importantly, mothers experiencing preeclampsia and gestational hypertension were pooled here, as the original authors did not distinguish between these nor was a subtype of preeclampsia reported.

### Identifying differentially expressed mitochondrial genes and interacting genes

To identify all mitochondrial differentially expressed genes (mtDEGs) present for maternal peripheral blood plasma samples, differential gene expression analysis was conducted using the DESeq2 package[41] available for R software[42]. Contrasts were first conducted for each collection timepoint (e.g., one contrast between normotensive and preeclamptic pregnancies for 1^st^ trimester, one contrast between normotensive and preeclamptic pregnancies for 2^nd^ trimester, etc.), where covariates significantly stratifying gene expression data at each timepoint were included in each timepoint DESeq2 contrast model. To do this, patient covariates were first investigated for autocorrelation and found infant birth weight to be correlated with placental weight, infant head circumference, and infant length (Supplementary Figure 1). Maternal age and body mass index (BMI) were separately correlated (Supplementary Figure 1). Correlated fetal and placental metrics were subsequently collapsed into a summary statistic by conducting principal component analysis (PCA) and taking the eigenvector associated for PC1 (>60% variance explained, data not shown) for each patient, and this process was also carried out on BMI and maternal age for each patient (>60% variance explained, data not shown). Covariates determined as significantly stratifying data were identified by conducting PCA on raw counts for each timepoint and using general linear models to determine covariate effects (main effects and interaction with preeclampsia diagnosis) on PC1 loadings. P-values ≤ 0.1 were used to allow for greater sensitivity in DESeq2 models (Supplementary Table S1). DEGs for each contrast were considered those with a P-adjusted value ≤ 0.05 (Supplementary File 1). An in-house DESeq analysis could not be conducted for fetal placental samples as patient characteristics were not provided in the original publication. We therefore relied on the DEG list provided by the authors (supplementary material of original publication[1]) to identify fetal DEGs. In these data, only P-adjusted value of significance and direction was provided. Therefore, to understand fetal mtDEG dynamics, the P-adjusted values for DEG list genes identified as mtDEGs were converted to Z-scores using one-tailed standard P-value distributions. We reported the lower tail for down-regulated gene expression and the upper tail for upregulated gene expression. Standard error was not provided by authors.

The MitoCarta 3.0 database[43], a database of all verified mitochondrial genes encoded by both the mitochondrial and nuclear genomes, was used to pull out mtDEGs by identifying mitochondrial gene matches to the MitoCarta database in all DEGs for both maternal and fetal samples (Supplementary Table S2). Once mtDEGs were identified, their respective interaction genes present in DEGs were found using String databases and an interaction confidence of “medium” or better (interaction score ≥ 0.400)[44]. For placental samples, interaction genes were pulled from the DEG list provided using the same interaction criteria cutoff (≥ 0.400). All possible interaction genes identified from String databases were used to conduct longitudinal gene set analyses for collections prior to delivery (1^st^ trimester, 2^nd^ trimester, and 3^rd^ trimester, details below).

### Maternal mitochondrial gene expression analysis during pregnancy

To investigate the functional consequences of aberrant maternal mitochondrial gene expression throughout pregnancy, Time Course Gene Set Analysis (TcGSA) was conducted using the TcGSA package (package version 0.12.6) available for R[45] and Ingenuity Pathway Analysis (IPA, QIAGEN Inc., https://www.qiagenbioinformatics.com/products/ingenuity-pathway-analysis). Delivery collection data were excluded from this analysis due to the unique physiology of labor and delivery. Transcripts counts were total count normalized[46] by counts per million. Outliers were identified and removed via Euclidean distance.

Default settings for TcGSA were used. DESeq was used to normalize gene counts across maternal gestation (covariates for DESeq model were chosen as described above). A custom gene matrix transposed (GMT) file (Supplementary File 2) was made using the interaction genes for each mtDEG identified during gestation, where gene sets were comprised of the interaction genes identified in String DB for each specified mtDEG. A null model was tested by making a second, size-matched GMT file comprised of non-mitochondrial and non-differentially expressed genes to ensure that significant gene sets identified in TcGSA were not an artifact of gene set size. Significant gene sets were considered those meeting a P-adjusted value ≤ 0.05 after TcGSA. Significant gene sets were probed for functional enrichment, details below.

### Functional enrichment analyses

Functional enrichments for mtDEG interaction genes present in maternal and fetal tissues were conducted using Gene Ontology enrichments (GO enrichment analysis[14]) and/or Ingenuity Pathway Analysis (IPA, QIAGEN Inc., https://www.qiagenbioinformatics.com/products/ingenuity-pathway-analysis). For maternal gestation, GO enrichment analyses were first conducted on individual significant gene sets using the Fisher’s exact test on the Gene Ontology web server and false discovery rate correction because the limited number of interaction genes present made IPA unfeasible. Full terms list for each gene set can be found in Supplementary File 2. All five of the significant TcGSA gene sets were primarily enriched for mitochondrial and metabolic homeostatic processes, while two of the five significant TcGSA gene sets (*MRPL38, FKBP8*) were additionally enriched for terms involved in immune system processes, developmental processes, and apoptotic processes (among others; Supplementary File 3). *MRPL38* and *FKBP8* gene sets were therefore combined to allow pathway enrichment in IPA for each gestational observation (1^st^ trimester, 2^nd^ trimester, and 3^rd^ trimester) using the Ingenuity Knowledge Base genes only reference set and specifying Human as the sample species. All other parameters followed default settings. Once the primary analysis was conducted, an additional comparison analysis was performed in IPA to determine pathway enrichment changes through time.

Due to the limited number of interaction genes present during delivery for both maternal and fetal tissues, IPA was not feasible. Therefore, to understand the functional consequences of aberrant mitochondrial gene expression in maternal and fetal tissues at delivery, GO enrichment analyses were conducted for the interaction genes using default settings on the Gene Ontology web server. Biological process and cellular component GO enrichments were performed and full terms list can be found in Supplementary File 3.

For plotting purposes, GO terms were matched to individual mtDEG interaction genes, and corresponding P-adjusted values for GO term enrichments were converted to Z-scores as described above, using the log-fold change of the mtDEG interaction gene to inform the direction of the Z-score (Supplementary File 4). Terms were then condensed using the “reduce_overlap” function in the GO Plot package[47] available for R, specifying a 99% overlap as the criteria for term collapsing.

## Supplementary Tables and Figures

**Supplementary Figure 1.**
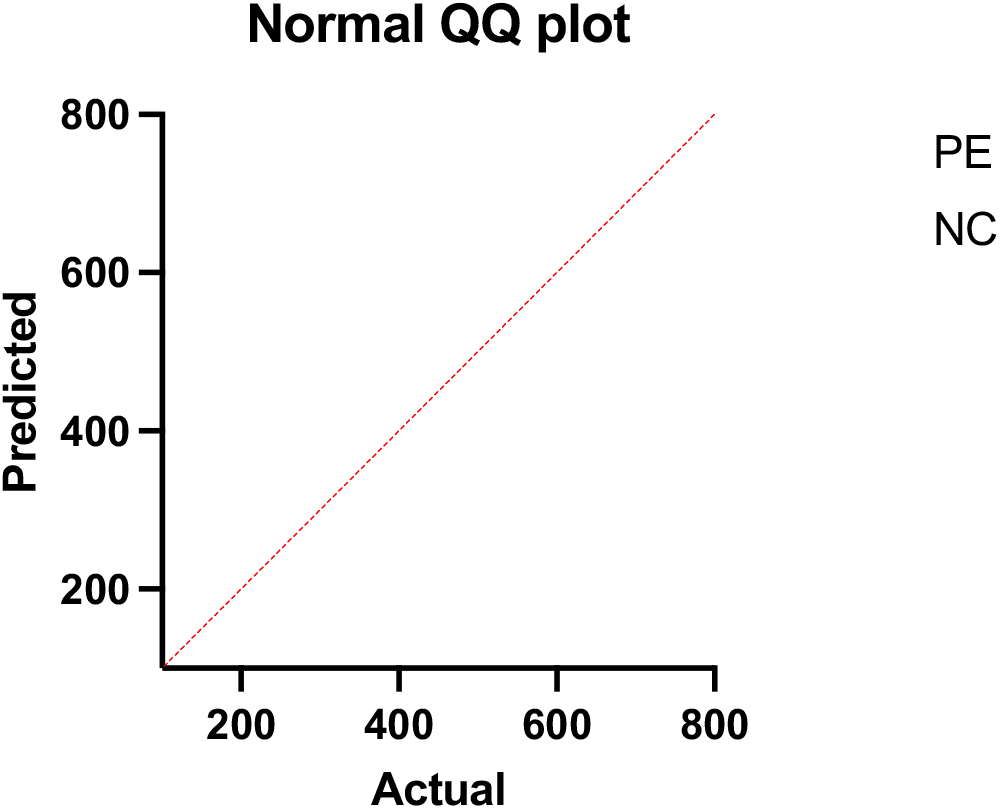
QQ plot of maternal mutational load of ccf-mtDNA

**Supplementary Figure 2.**
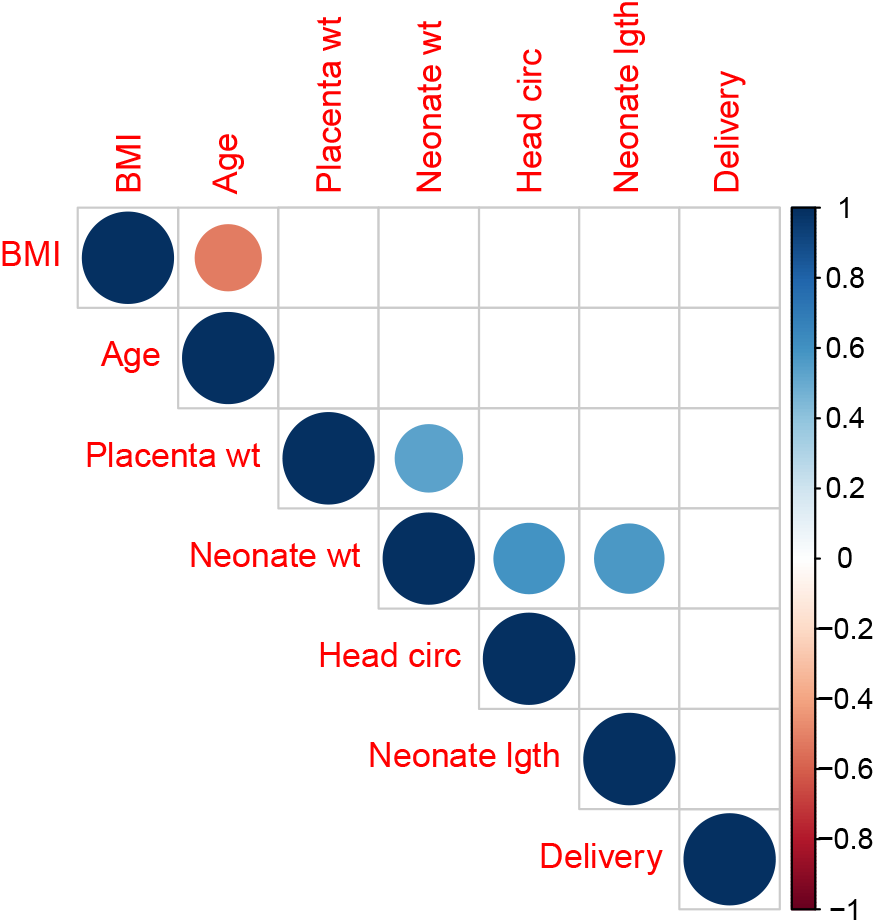
Correlation plot of maternal-fetal patient metrics for longitudinal study. Colored scale represents strength and direction of correlation; presence of circle indicates significant P-value (P-value ≤ 0.05); size of circle corresponds with significance, where larger circles represent smaller P-values; ‘BMI’: body mass index, ‘Age’: maternal age at delivery, ‘Placenta wt’: placental weight (g), ‘Neonate wt’: neonate weight at delivery (g), ‘Head circ’: neonate head circumference at delivery (cm), ‘Neonate lgth’: neonate length at delivery (cm), ‘Delivery’: neonate gestational age at delivery (weeks)

**Table S1.** Data structuring for maternal DESeq2 contrasts. PC1 eigen values for each collection were investigated for significant stratification by covariates using linear models. Covariates with a significant effect on PC1 eigen value stratification were included in DESeq model for respective collection contrasts (columns). P-value ≤ 0.1 used to allow for greater sensitivity in DESeq model.

| Covariate | 1<sup>st</sup> Trimester | 2<sup>nd</sup> Trimester | 3<sup>rd</sup> Trimester | Delivery |
| --- | --- | --- | --- | --- |
| Delivery mode | **P = 0.00616**<br>(main effect*)<br>**P = 0.03674**<br>(interactive effect*) | -- | -- | -- |
| Fetal sex | -- | -- | -- | -- |
| 1<sup>st</sup> Trimester collection (wks gestation) | -- | -- | -- | -- |
| 2<sup>nd</sup> Trimester collection (wks gestation) | -- | P = 0.0711<br>(main effect) | -- | -- |
| 3<sup>rd</sup> Trimester collection (wks gestation) | -- | -- | P = 0.0853<br>(main effect) | P = 0.084100<br>(interactive effect) |
| Delivery (wks gestation) | P = 0.0923 | -- | -- | **P = 0.046581**<br>(main effect)<br>**P = 0.018451**<br>(interactive effect) |
| Utero summary statistic | **P = 0.0298**<br>(main effect)<br>**P = 0.0500**<br>(interactive effect) | **P = 0.0142**<br>(main effect)<br>**P = 0.0271**<br>(interactive effect) | -- | -- |
| Age-BMI summary statistic | -- | -- | **P = 0.00708**<br>(main effect)<br>**P = 0.02846**<br>(interactive effect) | -- |

Values listed if P-value $\leq 0.1$ cut off was met. Values bolded if P-value $\leq 0.05$ ; (\*): specific to spontaneous delivery; 1<sup>st</sup> trimester total DEGs: 427; 2<sup>nd</sup> trimester total DEGs: 121; 3<sup>rd</sup> trimester total DEGs: 2345; Delivery total DEGs: 2148

**Table S2.** mtDEGs detected in maternal and fetal tissues. mtDEGs were identified via matching of DEGs to MitoCarta3.0 databases

| Gene name | Gene ID | Sample origin | Time detected | Chromosome |
| --- | --- | --- | --- | --- |
| PRORP | ENSG00000100890 | Maternal plasma | 1<sup>st</sup> trimester | chr14 |
| MRPL38 | ENSG00000204316 | Maternal plasma | 1<sup>st</sup> trimester,<br>delivery | chr17 |
| FKBP8 | ENSG00000105701 | Maternal plasma | 3<sup>rd</sup> trimester | chr19 |
| AGXT | ENSG00000172482 | Maternal plasma | 3<sup>rd</sup> trimester | chr2 |
| SLC25A41 | ENSG00000181240 | Maternal plasma | 3<sup>rd</sup> trimester | chr19 |
| NDUFB1 | ENSG00000183648 | Maternal plasma | 3<sup>rd</sup> trimester | chr14 |
| COX7B2 | ENSG00000170516 | Maternal plasma | 3<sup>rd</sup> trimester | chr4 |
| FARS2 | ENSG00000145982 | Maternal plasma | 3<sup>rd</sup> trimester | chr6 |
| LYRM2 | ENSG00000083099 | Maternal plasma | 3<sup>rd</sup> trimester | chr6 |
| BCL2L1 | ENSG00000171552 | Maternal plasma | 3<sup>rd</sup> trimester,<br>delivery | chr20 |
| ACSM2B | ENSG00000066813 | Maternal plasma | 3<sup>rd</sup> trimester | chr16 |
| CYP11B2 | ENSG00000179142 | Maternal plasma | 3<sup>rd</sup> trimester | chr8 |
| SLC25A20 | ENSG00000178537 | Maternal plasma | 3<sup>rd</sup> trimester | chr3 |
| TOMM20L | ENSG00000196860 | Maternal plasma | 3<sup>rd</sup> trimester | chr14 |
| ACOT11 | ENSG00000162390 | Maternal plasma | Delivery | chr1 |
| C15orf48 | ENSG00000166920 | Maternal plasma | Delivery | chr15 |
| ECHDC1 | ENSG00000093144 | Maternal plasma | Delivery | chr6 |
| GPX1 | ENSG00000233276 | Maternal plasma | Delivery | chr3 |
| NAT8L | ENSG00000185818 | Maternal plasma | Delivery | chr4 |
| FECH | ENSG00000066926 | Maternal plasma | Delivery | chr18 |
| ALAS2 | ENSG00000158578 | Maternal plasma | Delivery | chrX |
| NDUFAF3 | ENSG00000178057 | Maternal plasma | Delivery | chr3 |
| ISCA1 | ENSG00000135070 | Maternal plasma | Delivery | chr9 |
| DNAJC4 | ENSG00000110011 | Maternal plasma | Delivery | chr11 |
| ACSM4 | ENSG00000215009 | Maternal plasma | Delivery | chr12 |
| DMAC2 | ENSG00000105341 | Maternal plasma | Delivery | chr19 |
| VDAC3 | ENSG00000078668 | Maternal plasma | Delivery | chr8 |
| ALDH7A1 | ENSG00000164904 | Maternal plasma | Delivery | chr5 |
| ETFA | ENSG00000140374 | Maternal plasma | Delivery | chr15 |
| CARKD | Q8IW45 | Fetal placenta | Delivery | chr13 |
| PRDX6 | P30041 | Fetal placenta | Delivery | chr1 |
| AMACR | Q9UHK6 | Fetal placenta | Delivery | chr5 |
| EFHD1 | Q9BUP0 | Fetal placenta | Delivery | chr2 |
| PC | P11498 | Fetal placenta | Delivery | chr11 |
| SFXN3 | Q9BWM7 | Fetal placenta | Delivery | chr10 |
| RDH13 | Q8NBN7 | Fetal placenta | Delivery | chr19 |
| PDK2 | Q15119 | Fetal placenta | Delivery | chr17 |
| HADH | P40939 | Fetal placenta | Delivery | chr2 |

*Gene IDs are Ensembl identifiers for maternal tissues and Uniprot identifiers for fetal tissues. Chromosome location obtained from MitoCarta databases*

**Table S3:** Mitochondrial interaction gene sets analyzed by TcGSA. Bold values represent significant gene sets. No gene sets were significant in null-model testing

| mtDEG Gene Set | P-adj value | Size (no. genes) |
| --- | --- | --- |
| **MRPL38** | **5.30E-10** | **128** |
| **FKBP8** | **0.00188544** | **112** |
| **COX7B2** | **0.01640858** | **65** |
| **SLC25A41** | **0.01640858** | **21** |
| **NDUFB1** | **0.02218035** | **99** |
| FARS2 | 0.10744061 | 86 |
| TOMM20L | 1 | 52 |
| AGXT | 1 | 197 |
| SLC25A20 | 1 | 76 |
| CYP11B2 | 1 | 73 |
| LYRM2 | 1 | 23 |
| BCL2L1 | 1 | 440 |
| ACSM2B | 1 | 17 |
| PRORP | 1 | 63 |

*P-value adjusted for multiple comparisons. Bold text represents gene sets meeting the $p \leq 0.05$ significance threshold. Size represents number of mitochondrial interaction genes included in each gene set*

**Table S4.**
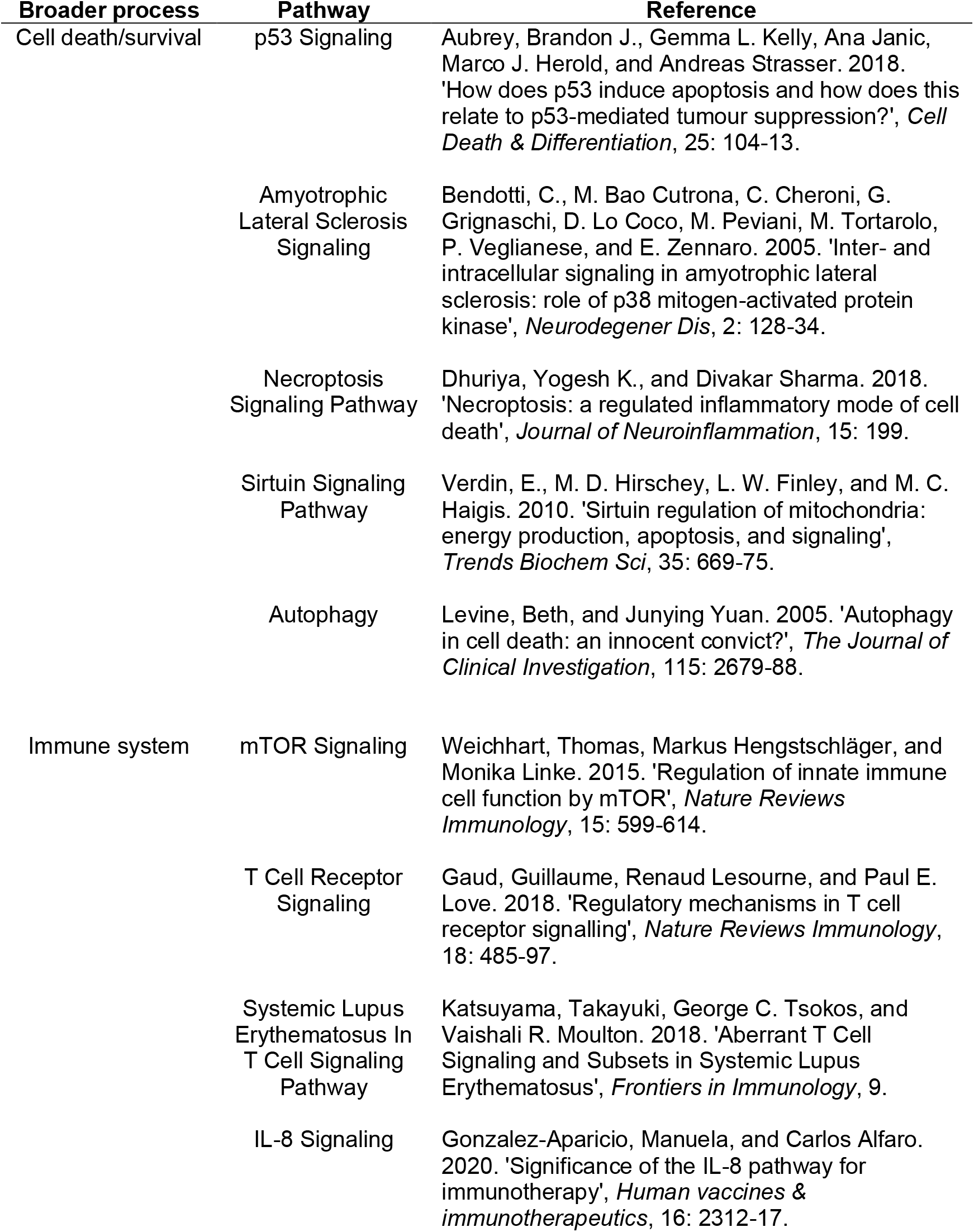
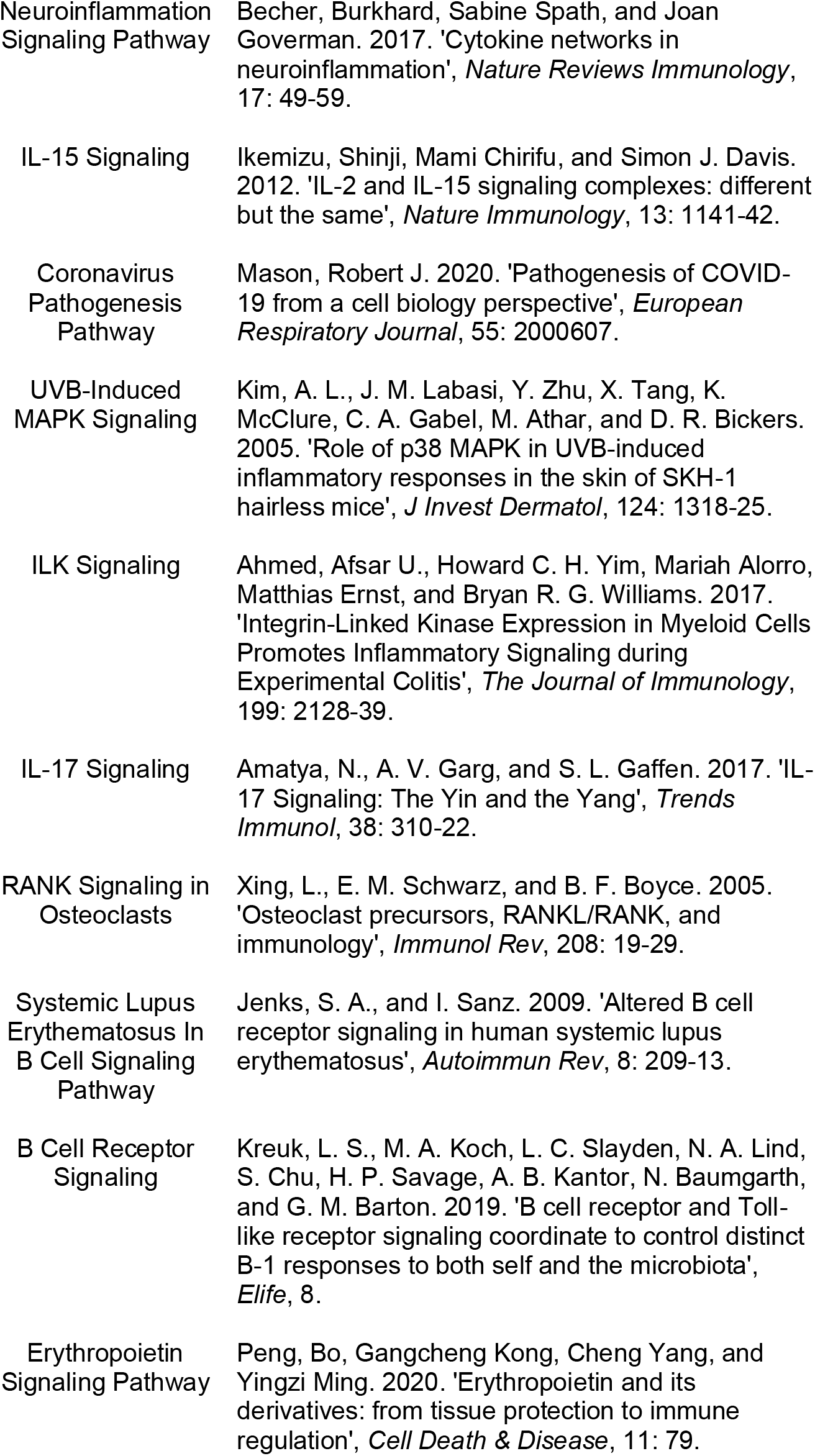
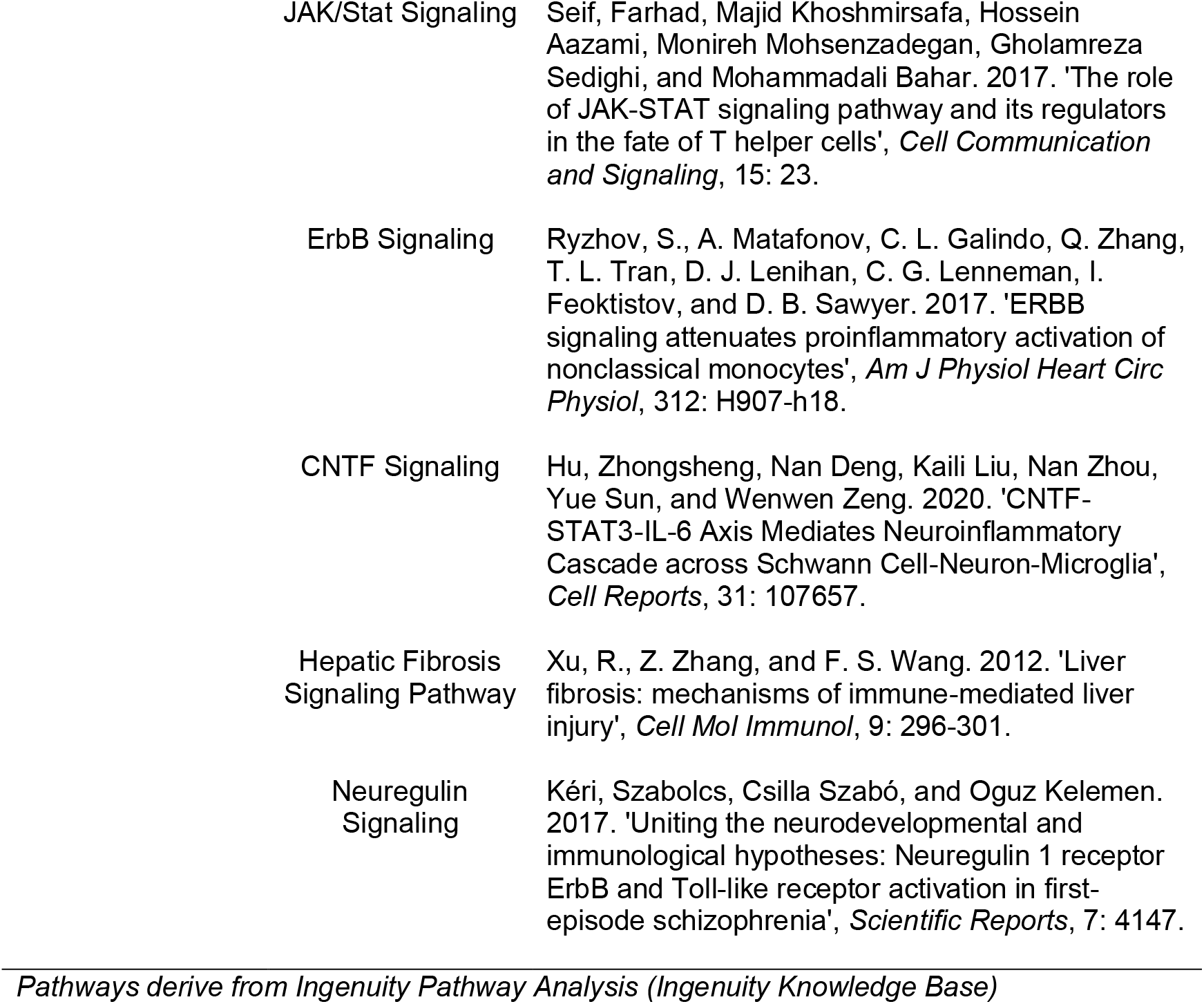
References supporting classification of *MRPL38* and *FKBP8* gene set pathway enrichments under the broader classifications of “Cell death/survival” and “Immune system”.

| Broader process | Pathway | Reference |
| --- | --- | --- |
| Cell death/survival | p53 Signaling | Aubrey, Brandon J., Gemma L. Kelly, Ana Janic, Marco J. Herold, and Andreas Strasser. 2018. 'How does p53 induce apoptosis and how does this relate to p53-mediated tumour suppression?', *Cell Death & Differentiation*, 25: 104-13. |
|  | Amyotrophic Lateral Sclerosis Signaling | Bendotti, C., M. Bao Cutrona, C. Cheroni, G. Grignaschi, D. Lo Coco, M. Peviani, M. Tortarolo, P. Veglianesi, and E. Zennaro. 2005. 'Inter- and intracellular signaling in amyotrophic lateral sclerosis: role of p38 mitogen-activated protein kinase', *Neurodegener Dis*, 2: 128-34. |
|  | Necroptosis Signaling Pathway | Dhuriya, Yogesh K., and Divakar Sharma. 2018. 'Necroptosis: a regulated inflammatory mode of cell death', *Journal of Neuroinflammation*, 15: 199. |
|  | Sirtuin Signaling Pathway | Verdin, E., M. D. Hirschey, L. W. Finley, and M. C. Haigis. 2010. 'Sirtuin regulation of mitochondria: energy production, apoptosis, and signaling', *Trends Biochem Sci*, 35: 669-75. |
|  | Autophagy | Levine, Beth, and Junying Yuan. 2005. 'Autophagy in cell death: an innocent convict?', *The Journal of Clinical Investigation*, 115: 2679-88. |
| Immune system | mTOR Signaling | Weichhart, Thomas, Markus Hengstschläger, and Monika Linke. 2015. 'Regulation of innate immune cell function by mTOR', *Nature Reviews Immunology*, 15: 599-614. |
|  | T Cell Receptor Signaling | Gaud, Guillaume, Renaud Lesourne, and Paul E. Love. 2018. 'Regulatory mechanisms in T cell receptor signalling', *Nature Reviews Immunology*, 18: 485-97. |
|  | Systemic Lupus Erythematosus In T Cell Signaling Pathway | Katsuyama, Takayuki, George C. Tsokos, and Vaishali R. Moulton. 2018. 'Aberrant T Cell Signaling and Subsets in Systemic Lupus Erythematosus', *Frontiers in Immunology*, 9. |
|  | IL-8 Signaling | Gonzalez-Aparicio, Manuela, and Carlos Alfaro. 2020. 'Significance of the IL-8 pathway for immunotherapy', *Human vaccines & immunotherapeutics*, 16: 2312-17. |

|  |  |
| --- | --- |
| Neuroinflammation Signaling Pathway | Becher, Burkhard, Sabine Spath, and Joan Goverman. 2017. 'Cytokine networks in neuroinflammation', *Nature Reviews Immunology*, 17: 49-59. |
| IL-15 Signaling | Ikemizu, Shinji, Mami Chirifu, and Simon J. Davis. 2012. 'IL-2 and IL-15 signaling complexes: different but the same', *Nature Immunology*, 13: 1141-42. |
| Coronavirus Pathogenesis Pathway | Mason, Robert J. 2020. 'Pathogenesis of COVID-19 from a cell biology perspective', *European Respiratory Journal*, 55: 2000607. |
| UVB-Induced MAPK Signaling | Kim, A. L., J. M. Labasi, Y. Zhu, X. Tang, K. McClure, C. A. Gabel, M. Athar, and D. R. Bickers. 2005. 'Role of p38 MAPK in UVB-induced inflammatory responses in the skin of SKH-1 hairless mice', *J Invest Dermatol*, 124: 1318-25. |
| ILK Signaling | Ahmed, Afsar U., Howard C. H. Yim, Mariah Alorro, Matthias Ernst, and Bryan R. G. Williams. 2017. 'Integrin-Linked Kinase Expression in Myeloid Cells Promotes Inflammatory Signaling during Experimental Colitis', *The Journal of Immunology*, 199: 2128-39. |
| IL-17 Signaling | Amatya, N., A. V. Garg, and S. L. Gaffen. 2017. 'IL-17 Signaling: The Yin and the Yang', *Trends Immunol*, 38: 310-22. |
| RANK Signaling in Osteoclasts | Xing, L., E. M. Schwarz, and B. F. Boyce. 2005. 'Osteoclast precursors, RANKL/RANK, and immunology', *Immunol Rev*, 208: 19-29. |
| Systemic Lupus Erythematosus In B Cell Signaling Pathway | Jenks, S. A., and I. Sanz. 2009. 'Altered B cell receptor signaling in human systemic lupus erythematosus', *Autoimmun Rev*, 8: 209-13. |
| B Cell Receptor Signaling | Kreuk, L. S., M. A. Koch, L. C. Slayden, N. A. Lind, S. Chu, H. P. Savage, A. B. Kantor, N. Baumgarth, and G. M. Barton. 2019. 'B cell receptor and Toll-like receptor signaling coordinate to control distinct B-1 responses to both self and the microbiota', *Elife*, 8. |
| Erythropoietin Signaling Pathway | Peng, Bo, Gangcheng Kong, Cheng Yang, and Yingzi Ming. 2020. 'Erythropoietin and its derivatives: from tissue protection to immune regulation', *Cell Death & Disease*, 11: 79. |
| JAK/Stat Signaling | Seif, Farhad, Majid Khoshmirsafa, Hossein Aazami, Monireh Mohsenzadegan, Gholamreza Sedighi, and Mohammadali Bahar. 2017. 'The role of JAK-STAT signaling pathway and its regulators in the fate of T helper cells', *Cell Communication and Signaling*, 15: 23. |
| ErbB Signaling | Ryzhov, S., A. Matafonov, C. L. Galindo, Q. Zhang, T. L. Tran, D. J. Lenihan, C. G. Lenneman, I. Feoktistov, and D. B. Sawyer. 2017. 'ERBB signaling attenuates proinflammatory activation of nonclassical monocytes', *Am J Physiol Heart Circ Physiol*, 312: H907-h18. |
| CNTF Signaling | Hu, Zhongsheng, Nan Deng, Kaili Liu, Nan Zhou, Yue Sun, and Wenwen Zeng. 2020. 'CNTF-STAT3-IL-6 Axis Mediates Neuroinflammatory Cascade across Schwann Cell-Neuron-Microglia', *Cell Reports*, 31: 107657. |
| Hepatic Fibrosis Signaling Pathway | Xu, R., Z. Zhang, and F. S. Wang. 2012. 'Liver fibrosis: mechanisms of immune-mediated liver injury', *Cell Mol Immunol*, 9: 296-301. |
| Neuregulin Signaling | Kéri, Szabolcs, Csilla Szabó, and Oguz Kelemen. 2017. 'Uniting the neurodevelopmental and immunological hypotheses: Neuregulin 1 receptor ErbB and Toll-like receptor activation in first-episode schizophrenia', *Scientific Reports*, 7: 4147. |

*Pathways derive from Ingenuity Pathway Analysis (Ingenuity Knowledge Base)*

## References

1. Awamleh, Z., G.B. Gloor, and V.K.M. Han, Placental microRNAs in pregnancies with early onset intrauterine growth restriction and preeclampsia: potential impact on gene expression and pathophysiology. BMC Medical Genomics, 2019. 12(1): p. 91.

2. Marín, R., et al., Oxidative stress and mitochondrial dysfunction in early-onset and late-onset preeclampsia. Biochim Biophys Acta Mol Basis Dis, 2020. 1866(12): p. 165961.

3. Vaka, V.R., et al., Role of Mitochondrial Dysfunction and Reactive Oxygen Species in Mediating Hypertension in the Reduced Uterine Perfusion Pressure Rat Model of Preeclampsia. Hypertension, 2018. 72(3): p. 703–711.

4. Cushen, S.C., et al., Concentrations and immunostimulatory potential of circulating cell-free membrane-bound and membrane-unbound mitochondrial DNA in preeclampsia. medRxiv, 2021: p. 2021.02.02.21250841.

5. Goulopoulou, S., et al., Toll-like receptor 9 activation: a novel mechanism linking placenta-derived mitochondrial DNA and vascular dysfunction in pre-eclampsia. Clin Sci (Lond), 2012. 123(7): p. 429–35.

6. Busnelli, A., et al., Mitochondrial DNA Copy Number in Peripheral Blood in the First Trimester of Pregnancy and Different Preeclampsia Clinical Phenotypes Development: A Pilot Study. Reprod Sci, 2019. 26(8): p. 1054–1061.

7. Del Vecchio, G., et al., Cell-free DNA Methylation and Transcriptomic Signature Prediction of Pregnancies with Adverse Outcomes. Epigenetics, 2021. 16(6): p. 642–661.

8. Hirashima, C., et al., Gestational hypertension as a subclinical preeclampsia in view of serum levels of angiogenesis-related factors. Hypertension Research, 2011. 34(2): p. 212–217.

9. Fernández-Silva, P., J.A. Enriquez, and J. Montoya, Replication and transcription of mammalian mitochondrial DNA. Exp Physiol, 2003. 88(1): p. 41–56.

10. Dorn, G.W., 2nd, R.B. Vega, and D.P. Kelly, Mitochondrial biogenesis and dynamics in the developing and diseased heart. Genes & development, 2015. 29(19): p. 1981–1991.

11. Yu, R., et al., Regulation of Mammalian Mitochondrial Dynamics: Opportunities and Challenges. Frontiers in Endocrinology, 2020. 11(374).

12. Costanzo, M.C. and T.D. Fox, Control of mitochondrial gene expression in Saccharomyces cerevisiae. Annu Rev Genet, 1990. 24: p. 91–113.

13. Clay Montier, L.L., J.J. Deng, and Y. Bai, Number matters: control of mammalian mitochondrial DNA copy number. J Genet Genomics, 2009. 36(3): p. 125–31.

14. Mi, H., et al., PANTHER version 14: more genomes, a new PANTHER GO-slim and improvements in enrichment analysis tools. Nucleic Acids Research, 2018. 47(D1): p. D419–D426.

15. Krämer, A., et al., Causal analysis approaches in Ingenuity Pathway Analysis. Bioinformatics, 2014. 30(4): p. 523–30.

16. Wallace, D.C., A mitochondrial paradigm of metabolic and degenerative diseases, aging, and cancer: a dawn for evolutionary medicine. Annu Rev Genet, 2005. 39: p. 359–407.

17. Cline, S.D., Mitochondrial DNA damage and its consequences for mitochondrial gene expression. Biochim Biophys Acta, 2012. 1819(9-10): p. 979–91.

18. Wallace, D.C., A mitochondrial bioenergetic etiology of disease. J Clin Invest, 2013. 123(4): p. 1405–12.

19. Zhang, Q., et al., Circulating mitochondrial DAMPs cause inflammatory responses to injury. Nature, 2010. 464(7285): p. 104–107.

20. Krysko, D.V., et al., Emerging role of damage-associated molecular patterns derived from mitochondria in inflammation. Trends Immunol, 2011. 32(4): p. 157–64.

21. Oka, T., et al., Mitochondrial DNA that escapes from autophagy causes inflammation and heart failure. Nature, 2012. 485(7397): p. 251–255.

22. McCarthy, F.P., et al., Evidence Implicating Peroxisome Proliferator-Activated Receptor- in the Pathogenesis of Preeclampsia. Hypertension, 2011. 58(5): p. 882–887.

23. Waite, L.L., R.E. Louie, and R.N. Taylor, Circulating Activators of Peroxisome Proliferator-Activated Receptors Are Reduced in Preeclamptic Pregnancy. The Journal of Clinical Endocrinology & Metabolism, 2005. 90(2): p. 620–626.

24. Geva, E., et al., Human placental vascular development: vasculogenic and angiogenic (branching and nonbranching) transformation is regulated by vascular endothelial growth factor-A, angiopoietin-1, and angiopoietin-2. J Clin Endocrinol Metab, 2002. 87(9): p. 4213–24.

25. Fan, X., et al., Endometrial VEGF induces placental sFLT1 and leads to pregnancy complications. The Journal of Clinical Investigation, 2014. 124(11): p. 4941–4952.

26. Cindrova-Davies, T., et al., Soluble FLT1 sensitizes endothelial cells to inflammatory cytokines by antagonizing VEGF receptor-mediated signalling. Cardiovascular Research, 2010. 89(3): p. 671–679.

27. Chau, K., A. Hennessy, and A. Makris, Placental growth factor and pre-eclampsia. J Hum Hypertens, 2017. 31(12): p. 782–786.

28. Maynard, S., F.H. Epstein, and S.A. Karumanchi, Preeclampsia and angiogenic imbalance. Annu Rev Med, 2008. 59: p. 61–78.

29. Pillay, P., et al., Placental exosomes and pre-eclampsia: Maternal circulating levels in normal pregnancies and, early and late onset pre-eclamptic pregnancies. Placenta, 2016. 46: p. 18–25.

30. Chang, X., et al., Exosomes From Women With Preeclampsia Induced Vascular Dysfunction by Delivering sFlt (Soluble Fms-Like Tyrosine Kinase)-1 and sEng (Soluble Endoglin) to Endothelial Cells. Hypertension, 2018. 72(6): p. 1381–1390.

31. Tannetta, D.S., et al., Characterisation of Syncytiotrophoblast Vesicles in Normal Pregnancy and Pre-Eclampsia: Expression of Flt-1 and Endoglin. PLOS ONE, 2013. 8(2): p. e56754.

32. Li, H., Aligning sequence reads, clone sequences and assembly contigs with BWA-MEM. arXiv preprint 1303.3997, 2013.

33. Danecek, P., et al., Twelve years of SAMtools and BCFtools. Gigascience, 2021. 10(2).

34. Van der Auwera, G.A. and B.D. O’Connor, Genomics in the cloud : using Docker, GATK, and WDL in Terra. 2020: O’Reilly Media, Inc.

35. Benjamin, D., et al., Calling Somatic SNVs and Indels with Mutect2. bioRxiv, 2019: p. 861054.

36. Cohen, J., A power primer. Psychol Bull, 1992. 112(1): p. 155–9.

37. Cohen, J., Statistical Power Analysis. Current Directions in Psychological Science, 1992. 1(3): p. 98–101.

38. Rice, M. and G. Harris, Comparing effect sizes in follow-up studies: ROCarea, Cohen’sd, andr. LawandHumanBehavior, 29,615–620. 2005.

39. Torchiano, M., Effsize - a package for efficient effect size computation. 2016: Zenodo.

40. Edgar, R., M. Domrachev, and A.E. Lash, Gene Expression Omnibus: NCBI gene expression and hybridization array data repository. Nucleic Acids Res, 2002. 30(1): p. 207–10.

41. Love, M.I., W. Huber, and S. Anders, Moderated estimation of fold change and dispersion for RNA-seq data with DESeq2. Genome Biology, 2014. 15(12): p. 550.

42. R Core Team, R: A Language and Environment for Statistical Computing. 2021, R Foundation for Statistical Computing: Vienna, Austria.

43. Rath, S., et al., MitoCarta3.0: an updated mitochondrial proteome now with sub-organelle localization and pathway annotations. Nucleic Acids Res, 2021. 49(D1): p. D1541–d1547.

44. Szklarczyk, D., et al., The STRING database in 2021: customizable protein– protein networks, and functional characterization of user-uploaded gene/measurement sets. Nucleic Acids Research, 2020. 49(D1): p. D605–D612.

45. Hejblum, B.P., J. Skinner, and R. Thiébaut, Time-Course Gene Set Analysis for Longitudinal Gene Expression Data. PLOS Computational Biology, 2015. 11(6): p. e1004310.

46. Evans, C., J. Hardin, and D.M. Stoebel, Selecting between-sample RNA-Seq normalization methods from the perspective of their assumptions. Briefings in Bioinformatics, 2017. 19(5): p. 776–792.

47. Walter, W., F. Sánchez-Cabo, and M. Ricote, GOplot: an R package for visually combining expression data with functional analysis. Bioinformatics, 2015. 31(17): p. 2912–4.

